# Impact of tiered restrictions on human activities and the epidemiology of the second wave of COVID-19 in Italy

**DOI:** 10.1101/2021.01.10.21249532

**Authors:** Mattia Manica, Giorgio Guzzetta, Flavia Riccardo, Antonio Valenti, Piero Poletti, Valentina Marziano, Filippo Trentini, Xanthi Andrianou, Alberto Mateo Urdiales, Martina del Manso, Massimo Fabiani, Maria Fenicia Vescio, Matteo Spuri, Daniele Petrone, Antonino Bella, Sergio Iavicoli, Marco Ajelli, Silvio Brusaferro, Patrizio Pezzotti, Stefano Merler

## Abstract

To counter the second COVID-19 wave in autumn 2020, the Italian government introduced a system of physical distancing measures organized in progressively restrictive tiers (coded as yellow, orange, and red) and imposed on a regional basis according to epidemiological risk assessments. The individuals’ attendance to locations outside the residential settings was progressively reduced with tiers, but less than during the national lockdown against the first COVID-19 wave in the spring. The reproduction number Rt decreased below the epidemic threshold in 85 out of 107 provinces after the introduction of the tier system, reaching average values of about 0.99, 0.89 and 0.77 in the yellow, orange and red tier, respectively. We estimate that the reduced transmissibility resulted in averting about 37% of the hospitalizations between November 5 and November 25, 2020. These results are instrumental to inform public health efforts aimed at preventing future resurgence of cases.

## Introduction

A second wave of COVID-19 has been spreading in all European countries in the fall of 2020^1^. In Italy, the daily incidence of confirmed cases rose slowly from 2 to 3 per 100,000 over the month of September, and then accelerated rapidly in October reaching a peak of 58 per 100,000 by November 13^1^. The second wave resulted in about 1.2 COVID-19-related deaths per 100,000 per day at the beginning of December; a value comparable to the first wave (1.35 per 100,000). The mortality rate has then declined to a stable plateau of about 0.8 deaths per 100,000 per day throughout the month of January^1, 2^. At the subnational level, a high geographical heterogeneity in the impact of the second wave was observed, with over four-fold variations across regions in the standardized mortality rate in October and November^3^.

To counter the rapid rise in SARS-CoV-2 infections observed since the end of September, the Italian government has progressively increased restrictions aimed at promoting physical distancing^4-7^. Between October 14 and November 5, 2020, interventions were uniformly enacted at the national level. These measures initially extended the mandatory use of face mask to outdoor spaces (previously mandated only indoors) and targeted a reduction of opening hours of bars and restaurants and a reduction of capacity of recreational venues such as cinemas and theaters. Shortly after, recreational venues and sports centers were closed altogether, and distance learning for at least 75% of the time was introduced in high schools. Starting from November 6, a three-tiered restriction system was introduced on a regional basis. A tier was assigned to each of the 21 regions and autonomous provinces (AP) by the Ministry of Health after an epidemiological risk assessment based on the combination of quantitative indicators on: i) the level of transmission, ii) the burden on older age groups and healthcare, and iii) public health resilience^7^. The sets of measures in the three tiers were labeled according to a color scheme: yellow, orange, and red, corresponding to increasing levels of restrictions. The tiered measures involved further limitations to retail and service activities, individual movement restrictions (ranging from a curfew between 10pm and 5am to a full-day stay-home mandate with a ban on inter-regional mobility), and reinforced distance learning in schools (see Table 1 for a complete list). The assignment of tiers to regions was updated according to frequent reassessments of the epidemiological situation.

**Table 1.** Description of restrictions applied in Italy since October 14^2-10^.

| Restrictions | October 14 – November 5, 2020 | November 6, 2020 – onwards |  |  |
| --- | --- | --- | --- | --- |
|  | National restrictions | Yellow tier | Orange tier | Red tier |
| **Face masks** | Mandatory in outdoor spaces | Mandatory in outdoor spaces | Mandatory in outdoor spaces | Mandatory in outdoor spaces |
| **Individual movements** | No restrictions | Stay-home mandate between 10pm and 5am (except for work, health and other certified reasons) | Stay-home mandate between 10pm and 5am and ban on movements between municipalities and to/from other regions (except for work, health and other certified reasons) | Stay-home mandate and ban on movements between municipalities and to/from other regions (except for work, health and other certified reasons). |
| **Retail and Services** | Open | Shopping malls closed during weekends and holidays (with the exception of essential retail & services) | Shopping malls closed during weekends and holidays (with the exception of essential retail & services) | All shops closed (with the exception of essential retail & services) |
| **Schools & Childcare** | Open until October 18. Recommendation to adopt distance learning for high schools and universities since October 19. Mandatory distance learning for at least 75% of the time in high schools since October 26.<br>Regional exceptions:<br>- in Campania, kindergartens were closed and distance learning for all schools adopted since October 16;<br>- in Apulia, distance learning for all schools adopted since October 30;<br>- in Lombardy, 100% distance learning for high schools since October 26;<br>- in Calabria, 100% distance learning for high schools and universities since October 26. | Distance learning in high schools and universities except when on-site attendance is essential (i.e., for laboratory activities) | Distance learning in high schools and universities except when on-site attendance is essential (i.e., for laboratory activities) | Distance learning in second and third grade of middle schools, in all grades of high schools and universities |
| **Bars serving food, Cafès & Restaurants** | No service after 12am until October 25. No service after 6pm and take away allowed until 12am since October 26. | No service after 6pm and take away allowed until 10pm. | Closed. Take away allowed until 10pm. | Closed. Take away allowed until 10pm. |
| **Public transport** | No capacity reduction<br>In Umbria, 50% capacity reduction since October 21. | 50% capacity reduction (except school service) | 50% capacity reduction (except school service) | 50% capacity reduction (except school service) |
| **Indoor recreational and cultural venues** | Open with capacity reduction until October 25. Closed since October 26. | Closed | Closed | Closed |
| **Gyms, pools & leisure venues** | Open until October 25. Non-professional contact sports not permitted. Closed except outdoor sport centers since October 26. | Closed except outdoor sport centers | Closed except outdoor sport centers | Individual outdoor training only (except sport events of national interest) |

The assessment of the effectiveness of the adopted interventions is critical to guide future decisions for the management of COVID-19 in Italy as well as other countries. In this work, we leverage the data from the Italian COVID-19 integrated surveillance system and publicly available mobility data^8^ to evaluate the impact of the three-tiered regional restriction system on human activities, SARS-CoV-2 transmissibility, hospitalization burden in Italy.

## Results

### Impact of tiered restrictions on human activities

The number of visitors in locations outside the residential settings decreased progressively increasing with the tier level (Figure 1 and Table 2). The most affected locations were those related to retail and recreation activities, as well as public transportation means, where, in the red tier, an over 50% reduction of presence was recorded with respect to pre-pandemic values (January 5 – February 6, 2020). The decline of attendance rates in these settings were mirrored by an increase of the time spent at home from 6.9% (95%CI: 4.2-10.6%) above the pre-pandemic values before the setup of tiered restrictions, to up to 17.7% (95%CI: 15.8-20.6%).

**Table 2.**
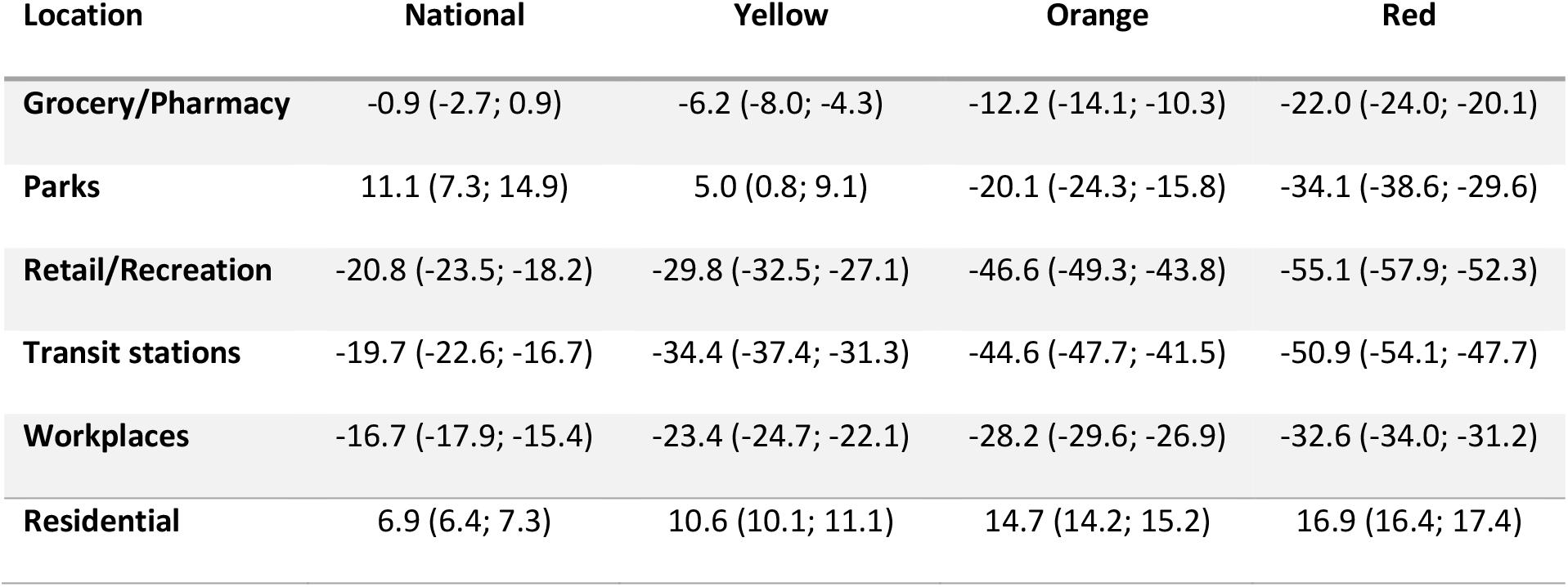
Change in the number of visitors in different locations relative to pre-pandemic values, as estimated by a linear mixed model (mean and 95%CI, values in percentage). For residential locations, the reported value refers to the changes in the time spent.

**Figure 1.**
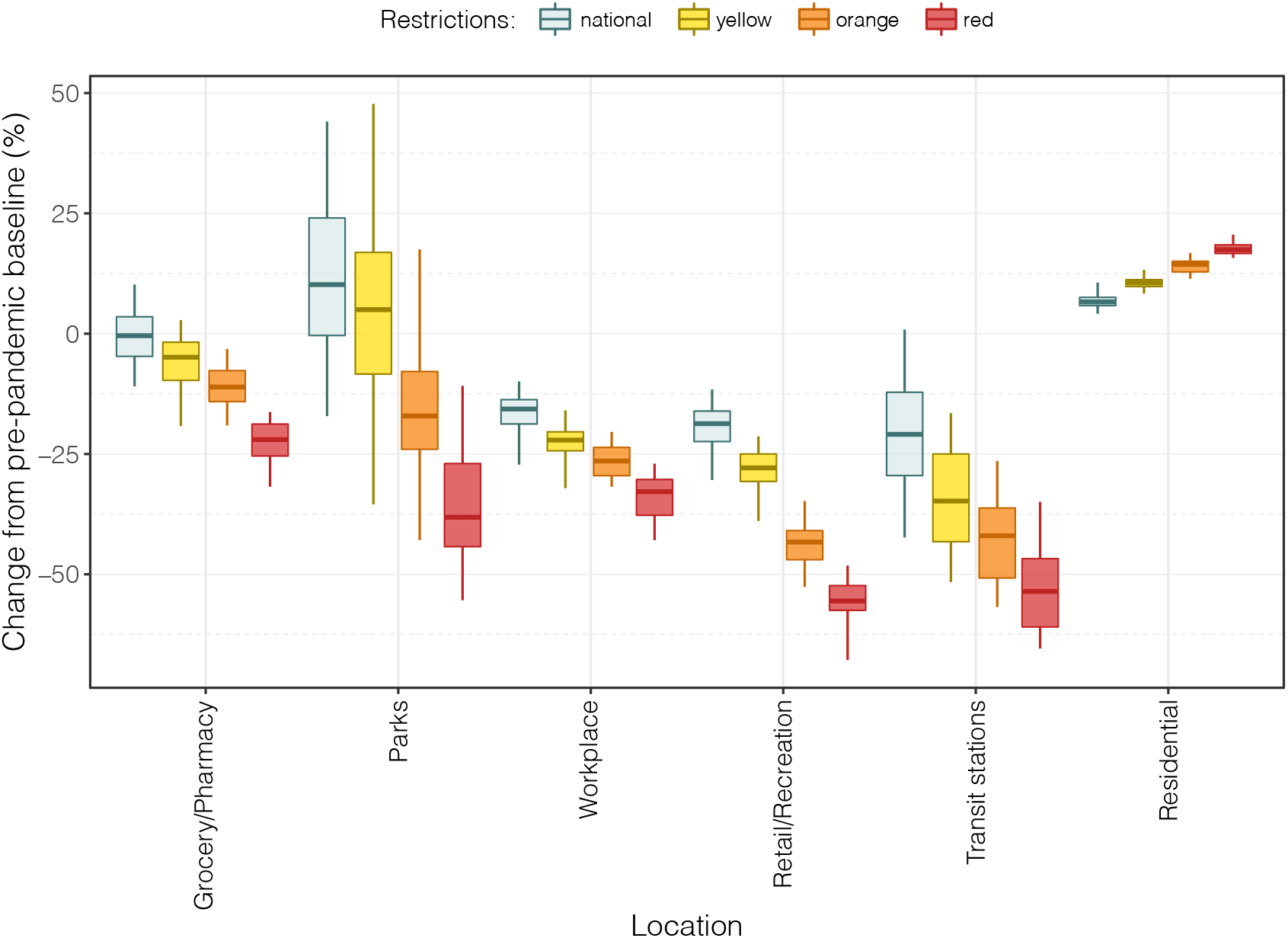
Distributions of changes in the time spent in different locations relative to pre-pandemic values, aggregated by tier (data at the province level^8^). Boxplots represent the median, interquartile range and 95% quantiles of the distributions.

### Impact of tiered restrictions on transmissibility

The temporal dynamics of the net reproduction number Rt in November 2020 were highly variable by region and province (Figure 2). For the purpose of this analysis, we group regions by the strictest tier assigned over the study period (November 6-25, 2020). In the week October 30-November 5, we estimated that the mean Rt for regions with maximum tier yellow, orange and red were 1.22 (95%CI 1.13-1.28), 1.35 (95%CI 1.17-1.53) and 1.40 (95%CI 1.21-1.53), respectively (Figure 3 and Appendix). These levels of SARS-CoV-2 transmissibility accounted for the imposition of national restrictions since October 14. We estimated that the introduction of the yellow tier resulted in a mean absolute Rt reduction of 0.22 (95%CI: 0.10 - 0.35) from the previous level. On top of this reduction, we estimated an additional reduction of 0.24 (95%CI: 0.09 - 0.39) for the orange tier, and of 0.40 (95%CI: 0.26 - 0.55) for the red tier (see Appendix). The net reproduction number between November 19 and 25 fell below the epidemic threshold in 42 of 46 (91%) provinces in the red tier, in 33 of 41 (81%) provinces in the orange tier and only in 10 of 20 provinces in the yellow tier (50%), despite the latter starting from much lower Rt values. These results were robust when aggregating the analysis at the regional, rather than provincial, level, when considering estimates of the reproduction number from hospitalized cases, when considering a different grouping of regions across tiers, and when using alternative lengths of the observation periods for Rt (see Appendix).

**Figure 2.**
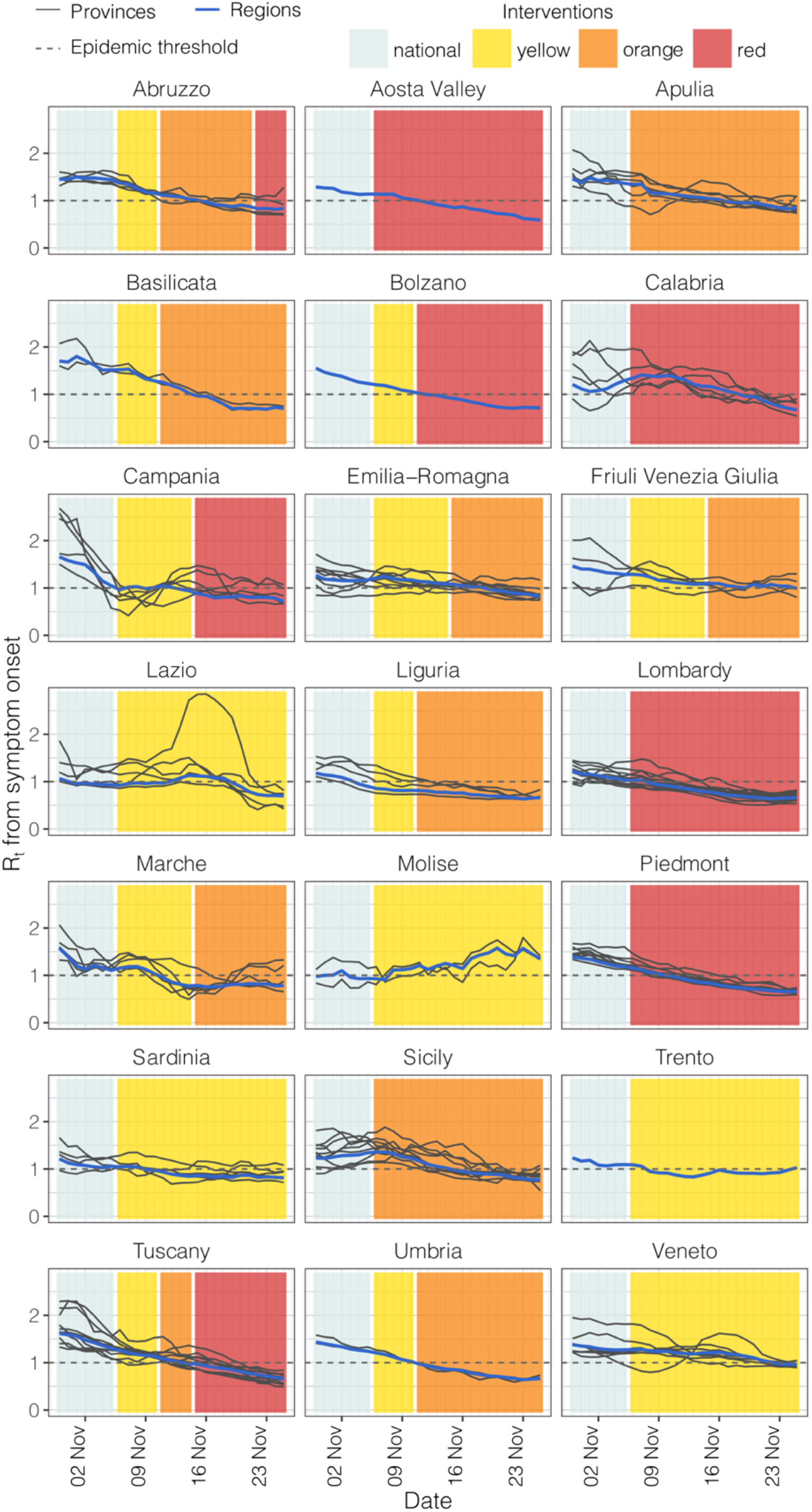
Temporal dynamics of the net reproduction numbers Rt and of the assigned tiers between October 30 and November 25. Each line shows the mean Rt for an Italian province (black) or region (blue). Provinces are grouped by region as tiers were assigned on a regional basis. Colored rectangles refer to the timeframe when the different tiers were adopted (see Table 1 for restrictions associated to the different tiers).

**Figure 3.**
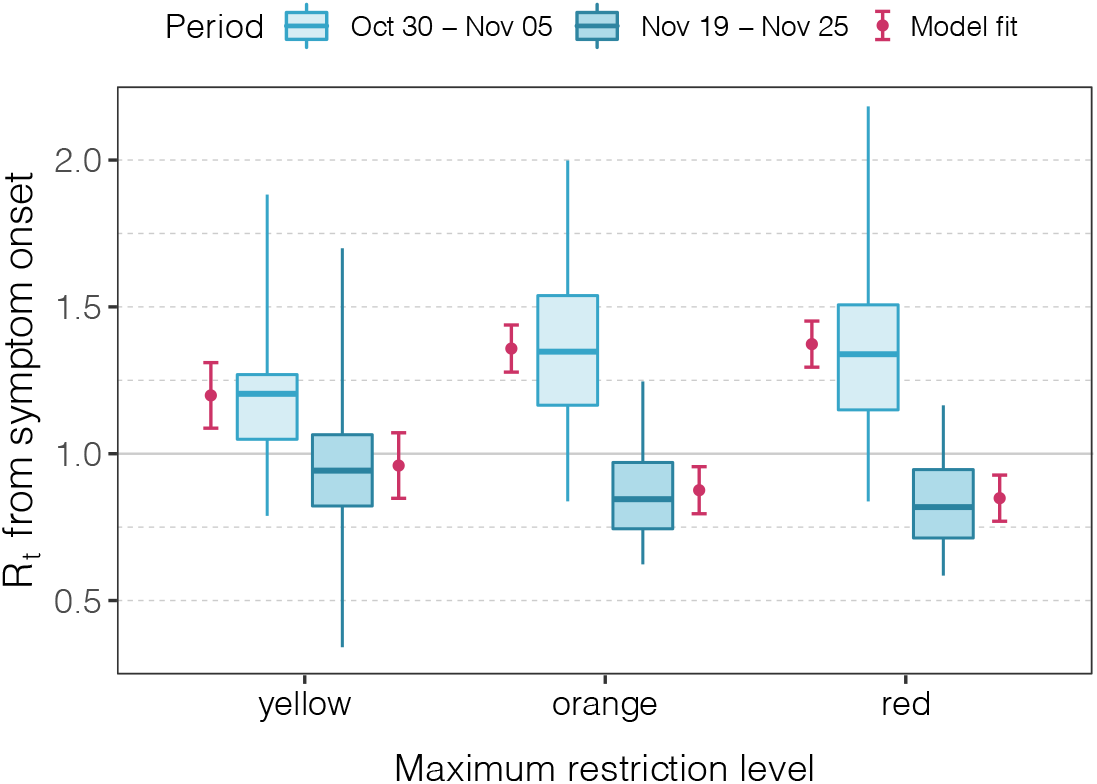
Distribution of the estimated reproduction numbers Rt across provinces aggregated according to tier and period of observation. Boxplots represent the median, interquartile range and 95% quantiles of the Rt distributions. Red dots represent the mean of the fit from the regression model and red vertical lines represent the 95% confidence interval around the mean.

### Impact of tiered restrictions on hospital admissions

We estimated the effect of imposing tiered restrictions on the cumulative hospital admissions over the study period (November 6 – 25, 2020) and on the daily hospitalization incidence at the end of the study period by comparing model outcomes under different sets of measures with respect to the observed values (Figure 4). The observed cumulative incidence of hospital admissions over the study period was 62.7, 66.3 and 83.8 per 100,000 in regions with maximum tier yellow, orange and red respectively, corresponding to 44,350 hospitalizations in the whole country. If the national restrictions that were in place on November 5 had been maintained until November 25, we estimate a cumulative incidence of hospital admissions of 75.6 (95%CI: 71.3 – 79.6), 107.4 (95%CI: 103.2 – 111.9) and 141.7 (95%CI: 137.8 – 145.9) per 100,000 in the yellow, orange and red tier respectively, resulting in a total of 70,063 (95%CI: 68,663-71,550) hospital admissions (Figure 4a). Thus, we estimate that the measures introduced by the government avoided about 36.7% (95%CI: 35.4-38%) of the overall hospitalizations in the considered period (i.e., slightly under 26,000), with a reduction of 16.9% (95%CI: 12.0-21.2%) in the yellow tier, 38.3% (95%CI: 35.7-40.7%) in the orange tier and 40.8% (95%CI: 39.2-42.5%) in the red tier. If regions in the yellow tier had been assigned to the orange or red tier, 14.7 (95%CI: 12.2-17.3) or 21.2 (95%CI: 19.0-23.3) hospital admissions per 100,000 could have been avoided respectively with respect to the observed values. If regions in the orange tier had been assigned to the red tier, 9.8 (95%CI: 7.8-11.9) hospital admissions per 100,000 could have been avoided with respect to the observed values.

**Figure 4.**
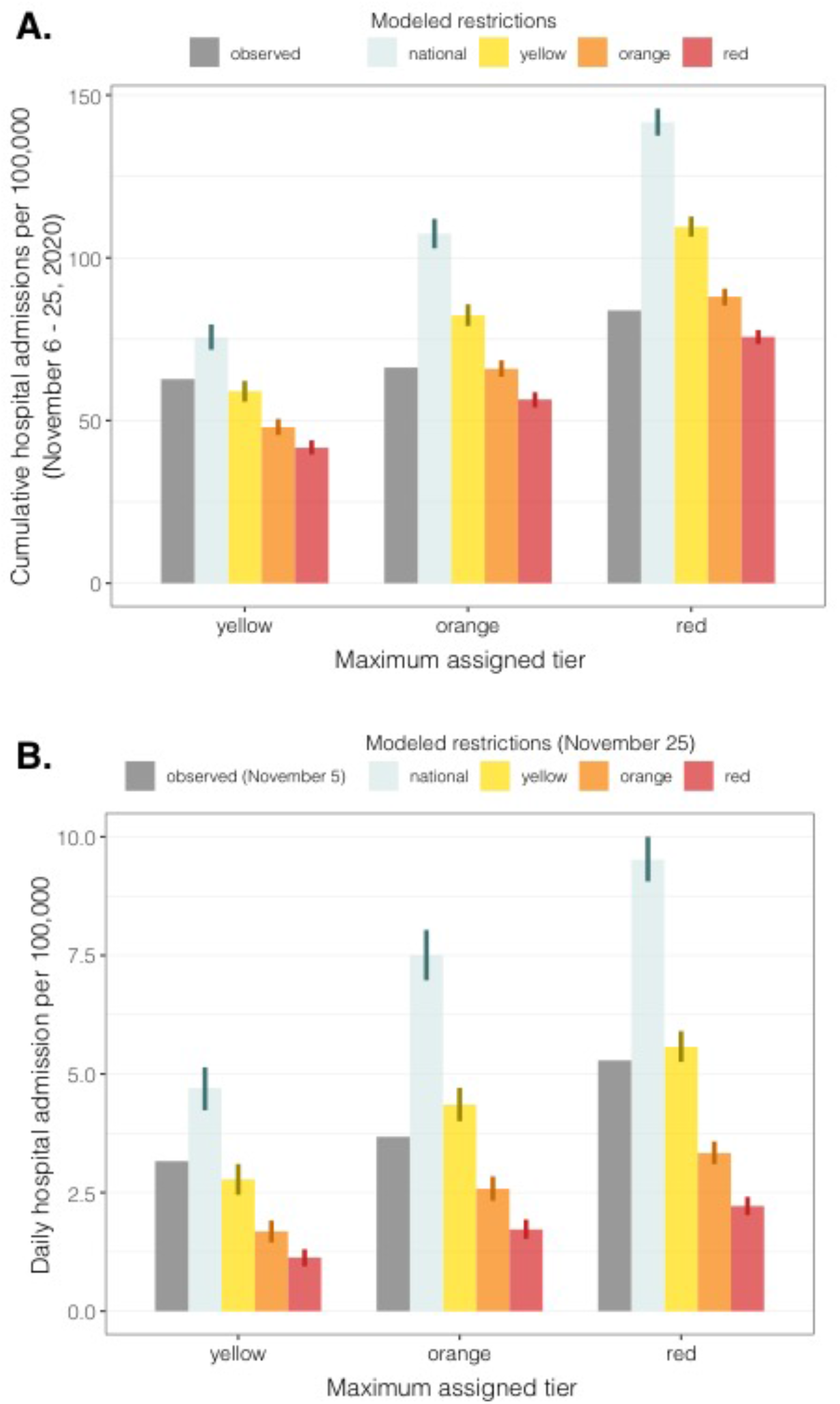
Incidence of hospital admissions by tier and modeled restrictions. A) Cumulative incidence over the projection period (November 6 – November 25, 2020). Gray bars represent the observed value for regions in different tiers. B) Incidence at the end of the projection period (November 25) in different scenarios. Grey bars represent the observed value at the beginning of the projection period (November 5) for regions in different tiers. Colored bars represent the mean projected value under the assumption that restrictions are maintained over the study period. Vertical lines represent the 95% confidence interval of the projected distribution.

If the tiered system had not been adopted, a general increase of the daily hospital admission incidence between November 6 and November 25 would have occurred; for example, in the red tier regions/APs the incidence would have been risen from 5.3 per 100,000 per day on November 5 to 9.5 (95%CI: 9.1-10.0) per 100,000 per day on November 25 (Figure 4b). The adoption of the yellow tier maintained roughly the same incidence level observed on November 5, while the orange and red tiers resulted in a consistent decline of the daily incidence.

## Discussion

The three-tiered restriction system introduced by the Italian government since November 6 on a regional basis has had a clear impact on both human activities and SARS-CoV-2 transmission. For what concerns human activities, we found a significant and progressive reduction of the time spent outside of home in all locations recorded by the Google mobility data^8^, especially those associated with recreational and retail activities and public transport. This is not surprising, considering that the restrictions mainly acted on social gathering venues that had not been targeted by previous interventions, such as bars, restaurants and shopping malls, and that they introduced limitations to individual movements. Reductions of attendance in schools (data not available in the Google mobility data) and workplaces may have contributed to the reduction in public transport use. However, we found that the activity reduction in all locations outside of home was far from that observed during the nation-wide lockdown imposed to counter the first wave, even in the strictest tier where a stay-home mandate was in place. As a comparison, during the lockdown the time spent in retail/recreational locations and in transit stations had dropped by over 75% with respect to the pre-pandemic values (against 50% in the red tier), and the time spent at home had increased by 27% (against 17% in the red tier)^8^.

On the epidemiological side, we found that the yellow tier reduced Rt to values close to 1, while orange and red tiers brought the reproduction number significantly below the epidemic threshold, even though starting from higher values. Overall, provinces in the yellow tier achieved a mean 18% reduction of Rt with respect to the transmission level determined by the preceding nationwide restrictions, while a reduction of 34% and 45% was observed in the orange and red tier respectively (Appendix). These results remained robust with respect to a number of sensitivity analyses, which considered a different type of data for the computation of Rt (hospitalized cases instead of symptomatic cases), a different level of geographic aggregation (regional instead of provincial), a different categorization of regions to account for changes over time in the assigned tier, and different windows of observations for Rt (Appendix). At the national level, the resulting reduction in transmissibility averted almost 26,000 hospital admissions between November 6 and November 25, 2020, with larger gains in regions assigned to stricter tiers. We note, however, that the estimated reduction in cumulative hospitalizations for the considered period does not fully represent the extent of the beneficial effects of the tier system. Indeed, the lower hospitalization incidence at the end of the projection period would extend its benefits on averted hospitalizations well after that date.

We acknowledge that during periods of high infection incidence there may be significant changes in notification rates due to the saturation of tracing and testing capabilities, and these changes may lead to biases in the estimates of the reproduction numbers^9,10^. During the second wave of COVID-19 in Italy, the largest increase in the number of cases occurred in October; therefore, we expect the notification rate to have stabilized before the period considered for Rt (October 30 – November 25). In addition, hospital admissions rates are less subject to changes compared with notification rates of symptomatic cases, and we found similar results when estimating transmissibility from hospital admissions. We also note that nationwide restrictions implemented to counter the second wave were scaled up in three different occasions (on October 14, 19 and 25^4-6^) before adopting the three-tiered regional system since November 6, 2020^7^. It is therefore possible that part of the decrease of Rt after November 6 is associated to a residual effect of earlier interventions. Previous studies have shown that most of the reduction in Rt takes place within about two weeks after the introduction of restrictions^11^. Therefore, this limitation should not have a major effect on our conclusions.

Our analysis is not suitable to pinpoint which specific restrictions maximized the reduction in transmissibility^12,13^, to disentangle the effect of spontaneous behavioral changes, and could not capture possible cross-regional effects. For example, provinces in the yellow tier sharing borders with regions in the orange or red tier may have indirectly benefited from a reduction of inter-regional mobility or that residents were more prone to self-imposing restrictions to their activity patterns.

## Conclusion

We quantified the epidemiological effect of the three-tiered system of restrictions adopted in Italy on a regional basis, showing that stricter restrictions (orange and red tiers) are needed to lower the incidence and that the most permissive tier (yellow) was sufficient to bring the reproduction number at approximately the epidemic threshold. We also showed that the tier system resulted in a much lower impact on human activities compared to lockdown. These insights are essential to support efforts to control the incidence of COVID-19 and to plan the response to possible future resurgence of cases, which could be also fueled by the emergence of more transmissible or virulent variants.

## Methods

### Restrictions data

We collected information from official sources on the measures taken by the Italian government between October 14^4-^7, 14^-18^ and November 25. Eleven of 21 regions and APs maintained the same tier from November 6 throughout the study period; for all remaining regions except Abruzzo, the highest tier corresponded also to the one which has been maintained for the longest time (see Figure 1 and Appendix).

### Mobility data

We retrieved data from the Google community mobility reports at the provincial level [8] over the period October 14 – November 25. These data represent the daily number of visitors at different locations, normalized to the value computed between January 5 and February 6, 2020 (pre-pandemic value). The locations reported in the data are categorized as follows: Grocery/Pharmacy (grocery markets, food warehouses, farmers markets, specialty food shops, drug stores, and pharmacies); Parks (local parks, national parks, public beaches, marinas, dog parks, plazas, and public gardens); Transit stations (public transport hubs such as subway, bus, and train stations); Retail/Recreation (restaurants, cafes, shopping centers, theme parks, museums, libraries, and movie theaters); and Workplaces (places of work). In addition, human activity in Residential (places of residence) is reported in terms of the mean duration of stay in these locations.

### Epidemiological data

Data to estimate the reproduction numbers and hospital admissions were collected by regional health authorities and collated by the Istituto Superiore di Sanità (Italian National Institute of Health) within an integrated surveillance system (described in ^19^). As a measure of transmissibility, we considered the net reproduction number Rt^9-^11, 20^^. The posterior distribution of Rt at any time point t was estimated by applying the Metropolis-Hastings MCMC sampling to a likelihood function defined as follows:

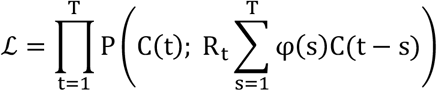

where

- P(k; λ) is the probability mass function of a Poisson distribution (i.e., the probability of observing k events if these events occur with rate λ).
- C(t) is the daily number of new cases having symptom onset at time t;
- R_t_ is the net reproduction number at time t to be estimated;
- φ(s) is the probability distribution density of the generation time evaluated at time s.

As a proxy for the distribution of the generation time, we used the distribution of the serial interval, estimated from the analysis of contact tracing data in Lombardy^21^, i.e., a gamma function with shape 1.87 and rate 0.28, having a mean of 6.6 days. We computed Rt for each of the 107 Italian provinces; as a sensitivity analyses, we computed Rt also for the 21 regions/APs and from the number of daily cases by date of hospital admission.

### Impact of tiered restrictions on human activities

In order to assess the impact of tiered restrictions on human activities, we applied the following linear mixed model to mobility data for each location category in the Google reports [8]:

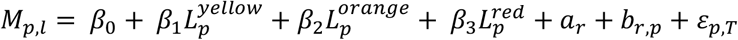

where

- *M*_*p,l*_ represents the mobility value (i.e., the change in the number of visitors in non-residential locations, or the change in the time spent at home, normalized to the pre-pandemic values) in each of the 107 Italian provinces (*p*), averaged over the days in which a given tier *l* was enforced;
- 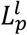 is a binary variable set to 1 when the considered value *Mp,l* belongs to a province with tier *l*, and 0 otherwise;
- *β*_0_, *β*_1_, *β*_2_ and *β*_3_ are model parameters, with *β*_0_ representing the mean mobility across Italian provinces during the period October 14-November 5 (i.e., before the tiered restriction system);
- *a*_*r*_ and *b*_*r,p*_ are random effects, assumed to be normally distributed. *a*_*r*_ allows random deviations from the mean among regions; *b*_*r,p*_ allows random deviations from the mean regional mobility among provinces within a region;
- *ε*_*p,T*_ is random noise assumed to be normally distributed.

### Impact of tiered restrictions on transmissibility

We applied a linear mixed model on the estimated SARS-CoV-2 transmissibility (i.e., the mean value of Rt):

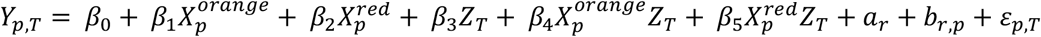

where

- *Y*_*p,T*_ represents the mean value of Rt in each of the 107 Italian provinces (*p*), averaged over two possible time periods (*T*): October 30 to November 5 (i.e., when nationwide interventions were still in place) or November 19 to November 25 (i.e., two to three weeks after the introduction of the tier system). In a sensitivity analysis, the length of the time periods is varied between 3 and 12 days (Appendix);
- 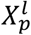 is a binary variable set to 1 if province *p* belongs to a region with maximum assigned tier *l*, and 0 otherwise;
- *Z*_*T*_ is a binary variable set to 0 if T=October 30 – November 5 and to 1 if T=November 19 – November 25;
- *β*_0_, *β*_1_, *β*_2_, *β*_3_, *β*_4_, and *β*_5_ are model parameters, with *β*_0_ representing the average value of Rt during the period October 30 – November 5 for provinces with maximum tier yellow;
- *a*_*r*_ and *b*_*r,p*_ are random effects, assumed to be normally distributed: *a*_/_ allows random deviations from the mean Rt among regions, *b*_*r,p*_ allows random deviations from the regional mean Rt among provinces within a region;
- *ε*_*p,T*_ is random noise assumed to be normally distributed.

### Impact of tiered restrictions on hospital admissions

For each region, we projected the curve of daily hospitalizations using the renewal equation^22^ under alternative assumptions on the implemented interventions. The number of new hospital admission H_i_(t) in a region *i* was given by

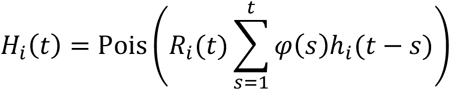

where

- *pois*(*λ*) is a Poisson sample with rate *λ*;
- h_i_(t) is the daily number of new hospital admissions in region *i*;
- *φ*(*s*) is the distribution of the generation time (see above);
- R_i_(t) is the assumed profile of the net reproduction number at time t in region *i*.

We used as input for h_i_(t) the curve of daily hospital admissions until November 5, 2020 (i.e., the day before the enactment of the tiered restrictions) and projected hospital admissions from November 6 to 25. To simulate the effect of maintaining restrictions of the pre-tier period versus the introduction of different tiers in each region, we considered different temporal profiles for R_i_(t). In all cases, the initial value of R_i_(t) on November 6 was taken as the mean value of Rt in the region, averaged between October 30 and November 5, 2020. Between November 19 and 25, we assume R_i_(t) to be constant and equal to a fraction *χ* = (1 − *ψ*) of the initial value, where *ψ* is the estimated mean relative reduction afforded by the tier with respect to the nationwide restrictions estimated by the linear model above (see Appendix). Between November 6 and 19, we assume a linear interpolation between the starting and final value. For nationwide restrictions, *ψ* = 0 by definition. This is equivalent to maintaining Rt constant and equal to the initial value throughout the projection period. We simulated 1,000 runs to accounting for stochastic variability of projections. To evaluate the effect of uncertainty around the estimation of R_i_(t), we carried out a sensitivity analysis by sampling, for each of the 1,000 stochastic simulations, the initial value of R_i_(t) at November 6 from the estimated posterior distribution of the mean value of Rt between October 30 and November 5 (Appendix).

## Data Availability

Data are available from the corresponding author upon reasonable request.

## Appendix

### 1. Impact of tiered restrictions on human activities

Table S1 reports the results of the linear mixed models on the Google mobility data for each of the location category. Parameter *β*_0_ represents the category’s mean mobility before the introduction of the tiers, while parameters *β*_1_, *β*_2_, *β*_3_ represents the difference in the mean mobility for provinces in yellow, orange and red tiers respectively, compared to *β*_0_. Reported values represent percentage points compared to the pre-pandemic baseline.

**Table S1.** Results of linear mixed models on Google mobility data for each location.

| Location | Parameter | Value | Std Error | DF | t-value | p-value |
| --- | --- | --- | --- | --- | --- | --- |
| Grocery/<br>Pharmacy | $\beta_0$ | -0.89925 | 0.917075 | -0.98056 | 15.94459 | 0.341454 |
| | $\beta_1$ | -5.25475 | 0.48196 | -10.9029 | 164.3782 | 3.59E-21 |
| | $\beta_2$ | -11.2807 | 0.517082 | -21.8162 | 164.9524 | 1.75E-50 |
| | $\beta_3$ | -21.1227 | 0.562332 | -37.5628 | 165.8015 | 5.29E-83 |
| Parks | $\beta_0$ | 11.09021 | 1.953271 | 5.677763 | 15.82006 | 3.58E-05 |
| | $\beta_1$ | -6.12371 | 1.433472 | -4.27194 | 166.9954 | 3.25E-05 |
| | $\beta_2$ | -31.1464 | 1.536525 | -20.2707 | 167.9877 | 5.38E-47 |
| | $\beta_3$ | -45.2031 | 1.668026 | -27.0997 | 169.4064 | 1.74E-63 |
| Residential | $\beta_0$ | 6.85952 | 0.237311 | 28.90514 | 21.68241 | 8.33E-19 |
| | $\beta_1$ | 3.709299 | 0.134483 | 27.58182 | 164.7453 | 1.23E-63 |
| | $\beta_2$ | 7.841163 | 0.14426 | 54.35424 | 165.4399 | 1.97E-107 |
| | $\beta_3$ | 10.07891 | 0.156845 | 64.26042 | 166.4686 | 1.98E-119 |
| Retail/<br>Recreation | $\beta_0$ | -20.8301 | 1.335306 | -15.5995 | 14.69864 | 1.50E-10 |
| | $\beta_1$ | -8.9815 | 0.534012 | -16.8189 | 165.9061 | 1.10E-37 |
| | $\beta_2$ | -25.7174 | 0.573161 | -44.8693 | 166.2494 | 8.09E-95 |
| | $\beta_3$ | -34.2708 | 0.623707 | -54.9469 | 166.7844 | 9.32E-109 |
| Transit stations | $\beta_0$ | -19.6636 | 1.511468 | -13.0096 | 16.93269 | 3.05E-10 |
| | $\beta_1$ | -14.725 | 0.6692 | -22.0039 | 160.1254 | 2.83E-50 |
| | $\beta_2$ | -24.9473 | 0.718081 | -34.7416 | 160.5276 | 1.41E-76 |
| | $\beta_3$ | -31.2401 | 0.787786 | -39.6556 | 161.8096 | 2.98E-85 |
| Workplaces | $\beta_0$ | -16.6515 | 0.655163 | -25.4159 | 19.38602 | 2.37E-16 |
| | $\beta_1$ | -6.74687 | 0.302839 | -22.2787 | 163.3531 | 2.20E-51 |
| | $\beta_2$ | -11.5679 | 0.324983 | -35.5954 | 163.7966 | 5.38E-79 |
| | $\beta_3$ | -15.9586 | 0.353539 | -45.1396 | 164.4871 | 1.40E-94 |

### 2. Impact on transmissibility

Figure S1 reports the temporal change observed in the estimated net reproduction number Rt across different provinces of Italy, after the introduction of regional tiered restrictions. Table S2 shows the aggregate estimate of Rt, before and after the introduction of tiered restrictions. Table S3 displays the results obtained with the linear mixed model applied to values of Rt at provincial level, according to our main analysis. Residuals obtained from this analysis are reported in Figure S2.

**Table S2.**
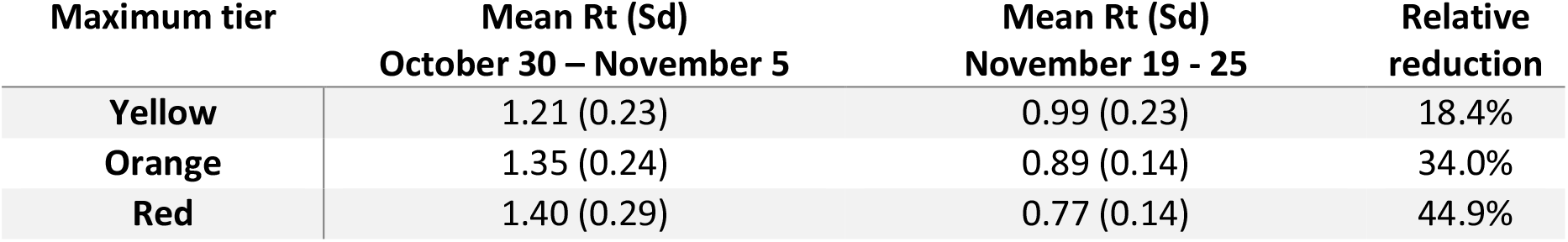
Net reproduction number Rt before and after the tiered restrictions (main analysis).

| Maximum tier | Mean $R_t$ (Sd)<br>October 30 – November 5 | Mean $R_t$ (Sd)<br>November 19 - 25 | Relative<br>reduction |
| --- | --- | --- | --- |
| Yellow | 1.21 (0.23) | 0.99 (0.23) | 18.4% |
| Orange | 1.35 (0.24) | 0.89 (0.14) | 34.0% |
| Red | 1.40 (0.29) | 0.77 (0.14) | 44.9% |

**Table S3.** Results of the linear mixed model on the net reproduction number Rt at provincial level (main analysis). The estimated standard deviation for the random effect between regions, *a*_*r*_, was 0.09, while the one for the random effect between provinces of the same region, *b*_*p,r*_ was 7·10^−10^. The estimated standard deviation for random noise, *ε*_*p,T*_, was 0.20.

| PARAMETER | INTERPRETATION | VALUE | STD<br>ERROR | DF | T-VALUE | P-VALUE |
| --- | --- | --- | --- | --- | --- | --- |
| $\beta_0$ | Mean $R_t$ before the introduction of tiers for provinces in maximum tier yellow | 1.215 | 0.061 | 37.8 | 18.830 | <0.00001 |
| $\beta_1$ | Difference in the mean $R_t$ before the introduction of tiers for provinces in maximum tier orange, compared to yellow | 0.138 | 0.076 | 35.3 | 1.821 | 0.07708 |
| $\beta_2$ | Difference in the mean $R_t$ before the introduction of tiers for provinces in maximum tier red, compared to yellow | 0.198 | 0.076 | 33.6 | 2.608 | 0.01348 |
| $\beta_3$ | Reduction in $R_t$ for provinces in maximum tier yellow | -0.224 | 0.063 | 193.2 | -3.559 | 0.00047 |
| $\beta_4$ | Additional reduction in $R_t$ for provinces in maximum tier orange, on top of reduction afforded by yellow | -0.235 | 0.077 | 193.2 | -3.063 | 0.00251 |
| $\beta_5$ | Additional reduction in $R_t$ for provinces in maximum tier red, on top of reduction afforded by yellow | -0.403 | 0.076 | 193.2 | -5.341 | <0.00001 |

**Figure S1.**
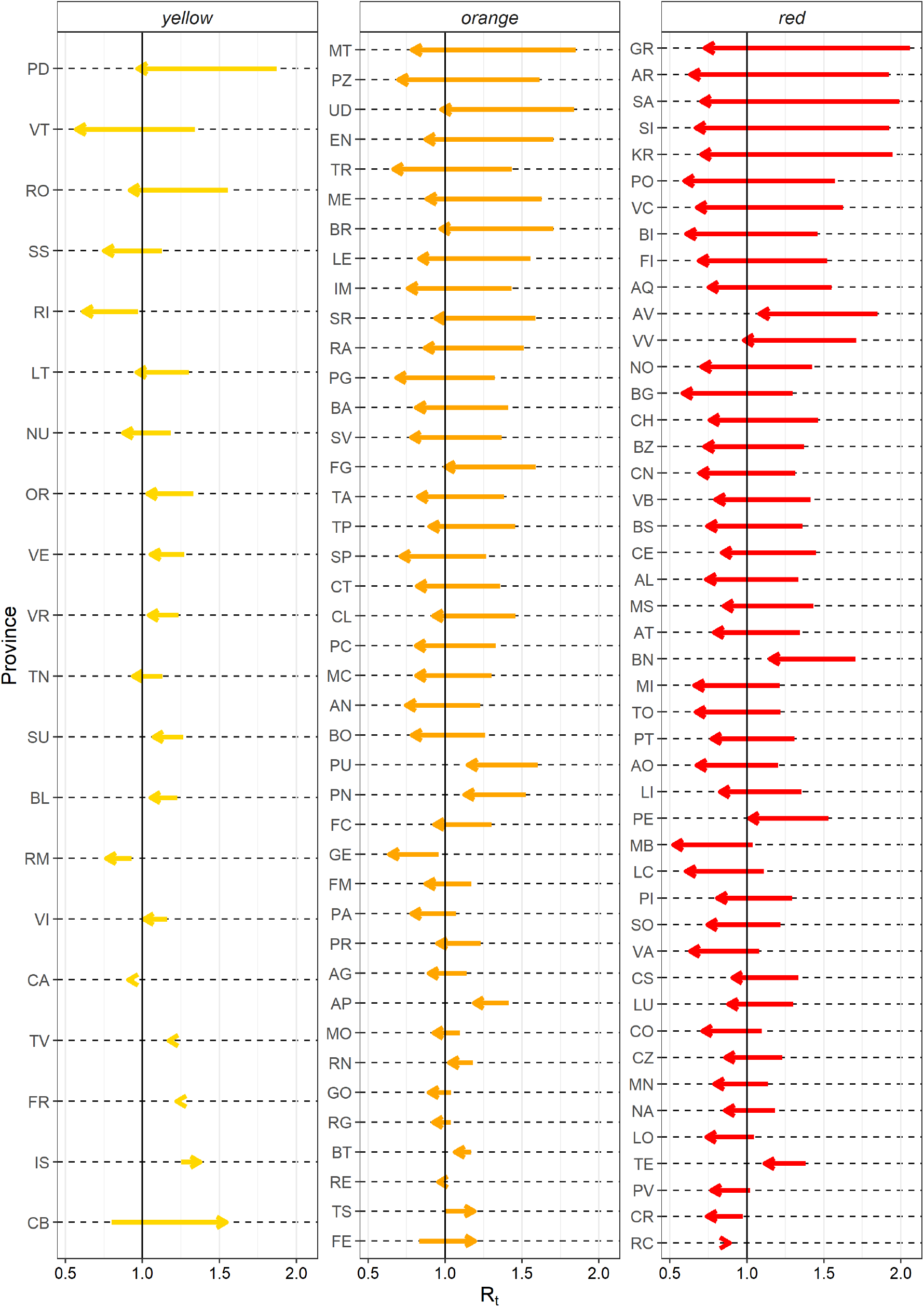
Variation of the net reproduction number Rt in each province. The arrows indicate the variation in Rt from the week before the introduction of regional tiered restrictions (October 30-November 5) to the end of our observations (November 19 – 25). Provinces are ordered by decreasing reduction in Rt.

**Figure S2.**
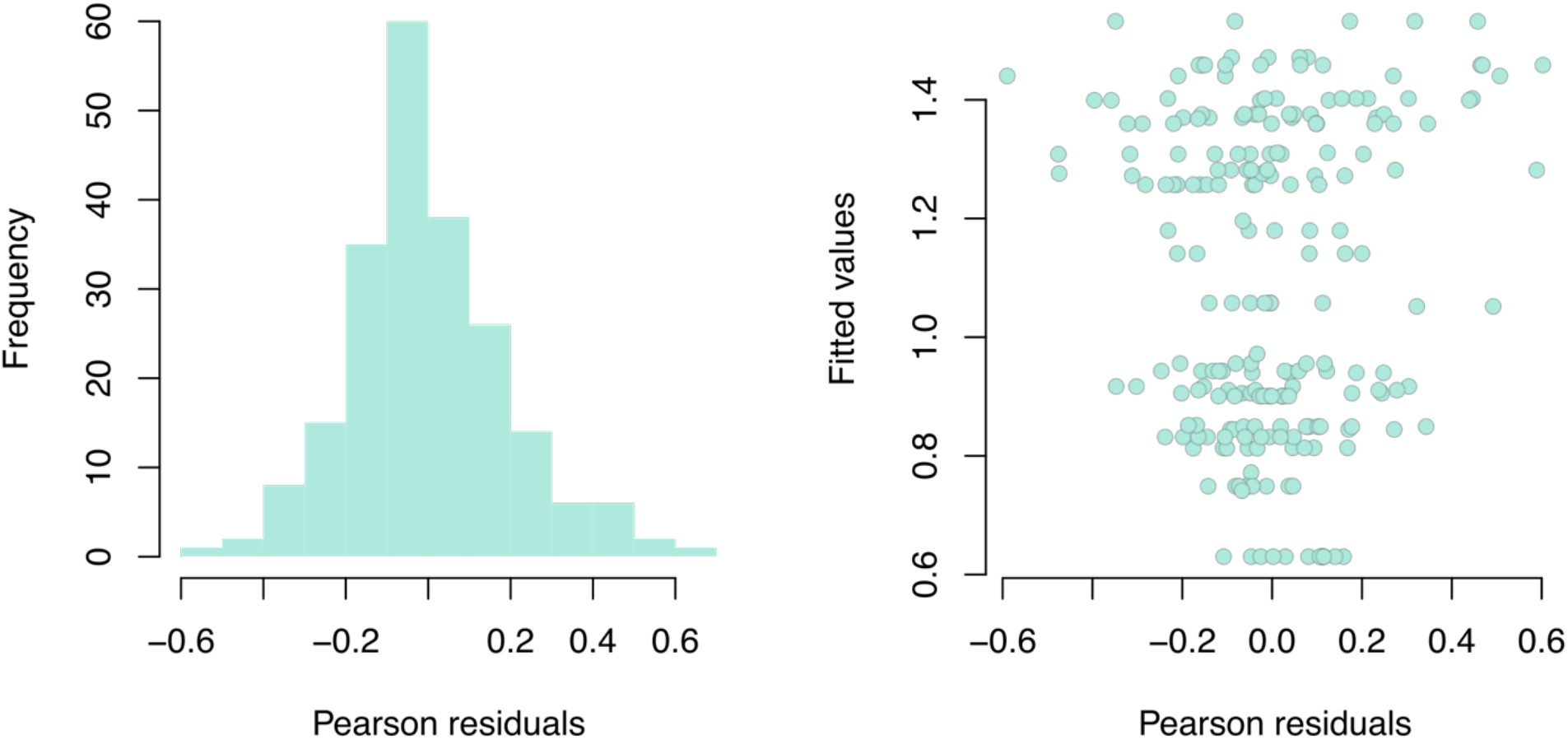
Analysis of residuals for the linear model (main analysis). Left: distribution of Pearson residuals (i.e., raw residuals normalized with respect to the variance of residuals); right: scatterplot between Pearson residuals and fitted values of *Y*_*p,T*_.

#### 2.1 Sensitivity analyses on transmissibility

We evaluated the robustness of our results by re-estimating the impact of tiered restrictions on transmissibility with alternative modelling assumptions. In particular, we considered the following four sensitivity analyses:

##### Hospital admission

We used the same model described in the main text, but considering as dependent variable Y_p,T_ the reproduction number from hospital admissions, Rt^h^, stratified by province and period of observation. Figure S3 shows the temporal change observed in the estimated Rt^h^ across different provinces of Italy, before and after the introduction of regional tiered restrictions. Results obtained with this sensitivity analysis are reported in Table S4 and S5, and Figure S4. In particular, Table S5 shows that the starting values of Rt^h^ (Oct 30 – Nov 5) were more homogeneous across tiers than those of Rt computed from symptomatic cases. However, estimated reductions associated with different tiers are comparable to those obtained in the main analysis.

##### Regional analysis

We applied the baseline regression model to region-specific data as follows:

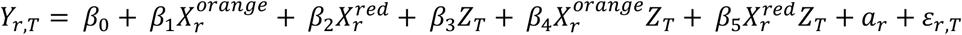

where Y_r,T_ represents the regional values of Rt, and where the independent variables are region-specific rather than province-specific. Obtained results are displayed in Tables S6 and S7. Due to the lower number of observations, model results for Rt show a lower statistical power compared with the baseline analysis. However, the estimated reductions associated with different tiers are comparable to those obtained in the main analysis.

##### Lengths of periods T

We applied the baseline regression model to Rt values computed at provincial level but considering average values over alternative period lengths ranging from 3 to 11 days. For example, when considering a period length of 3 days, we compared the mean Rt in period November 3 – 5 against November 23-25. Figure S5 shows the difference between model parameters estimated for each period length compared to the main analysis (7 days). Estimated variations in parameter estimates are limited (within 0.05 in most cases).

##### Alternative grouping of provinces

We applied the baseline regression model to Rt values computed at provincial level but considering a finer grouping of provinces with 5 categories rather than 3, to account for the temporal evolution of tier assignments to the region of belonging of the province. Specifically, we categorized tiers in five groups (see Figure 2 in the main text for a reference of category assignments):

- L1: tier constantly yellow (20 provinces): Lazio (5 provinces), Molise (2 provinces), Sardinia (5 provinces), Trento (1 Autonomous Province), Veneto (7 provinces);
- L2: tier reaching up to orange (26 provinces): Basilicata (2 provinces), Emilia-Romagna (9 provinces), Friuli Venezia Giulia (4 provinces), Liguria (4 provinces), Marche (5 provinces), Umbria (2 provinces);
- L3: tier constantly orange (15 provinces): Apulia (6 provinces), Sicily (9 provinces)
- L4: tier reaching up to red (20 provinces): Abruzzo (4 provinces), Bolzano (1 Autonomous Province), Campania (5 provinces), Tuscany (10 provinces);
- L5: tier constantly red (26 provinces): Aosta Valley (1 province), Calabria (5 provinces), Lombardy (12 provinces), Piedmont (8 provinces).

The obtained results were substantially equivalent to those presented in the main text (see Table S8 and S9).

**Table S4.**
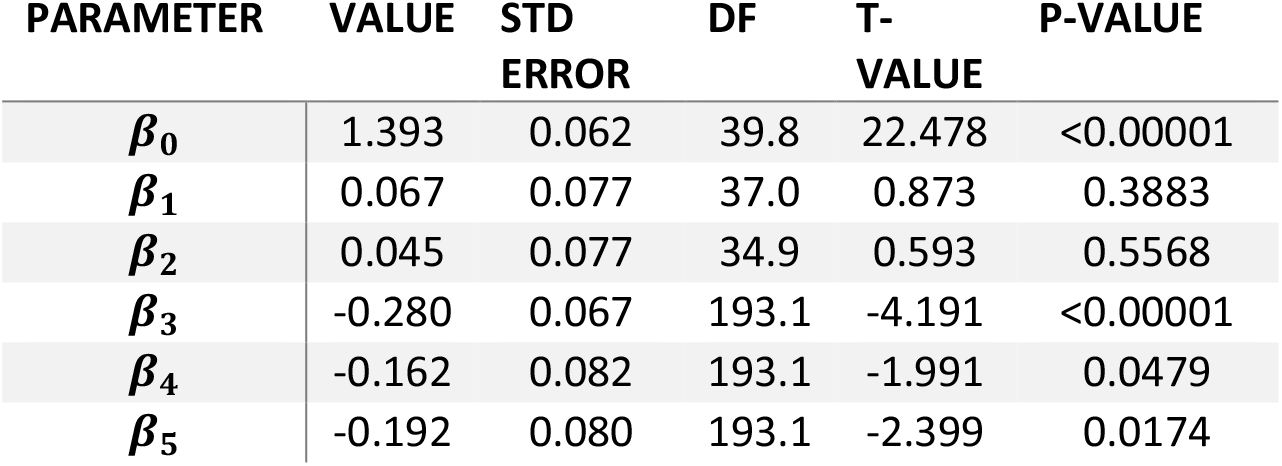
Result of the linear mixed model on the net reproduction number estimated from hospital admissions, Rt^h^, at the provincial level. See Table S2 for interpretation of parameters.

**Table S5.** Mean net reproduction number estimated from hospital admission, Rt^h^, before and after regional interventions.

| Maximum tier | Mean $R_t^h$ (SD)<br>October 30 – November 5 | Mean $R_t^h$ (SD)<br>November 19 - 25 | Relative<br>reduction |
| --- | --- | --- | --- |
| Yellow | 1.38 (0.21) | 1.10 (0.14) | 20.3% |
| Orange | 1.48 (0.26) | 1.03 (0.19) | 30.4% |
| Red | 1.44 (0.27) | 0.97 (0.20) | 32.6% |

**Figure S3.**
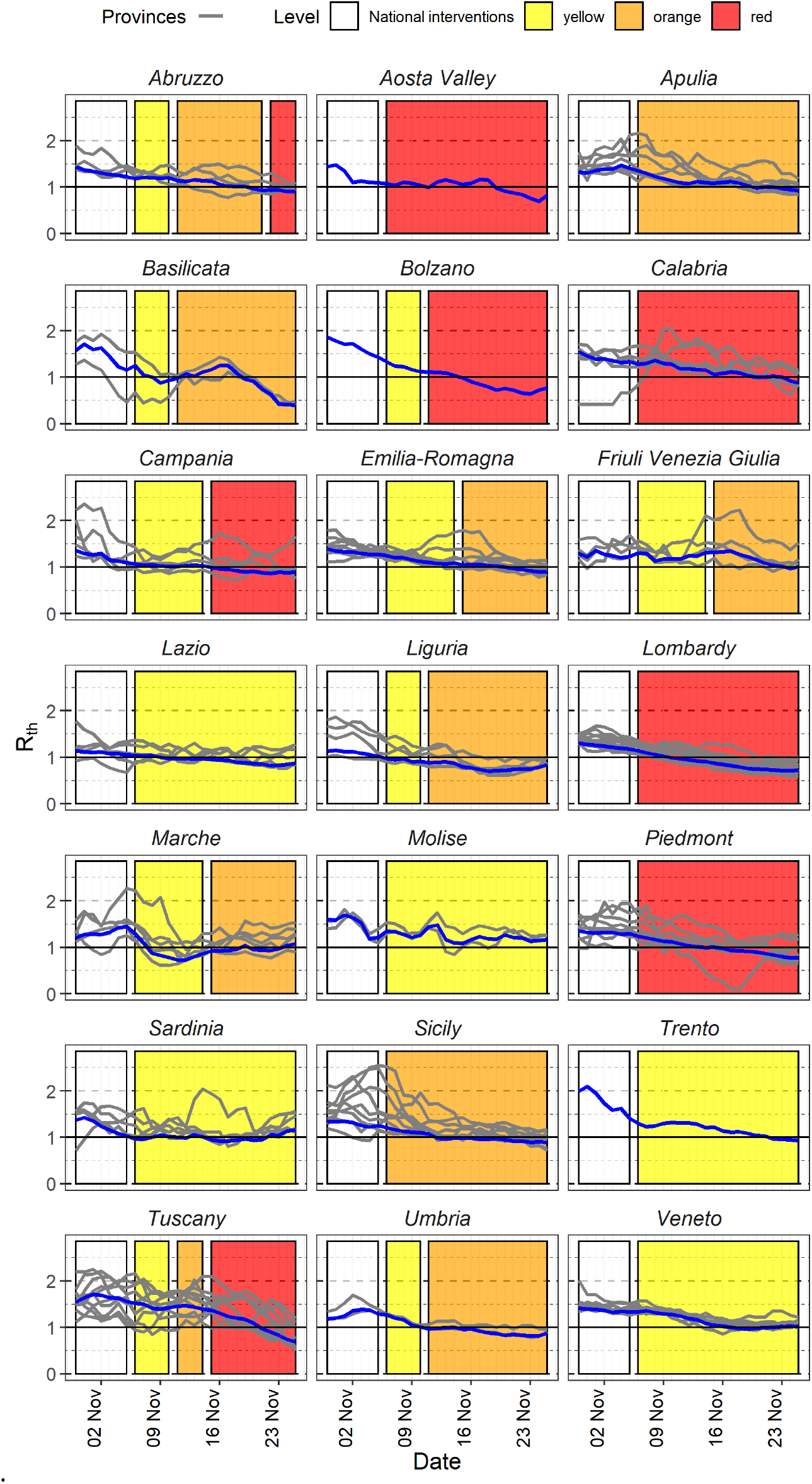
Temporal dynamics of the net reproduction numbers from hospital admissions, Rt^h^, and of restrictions applied between October 30 and November 25. Each line shows the mean Rt^h^ for an Italian province (grey) or region (blue). Provinces are grouped by region as interventions were carried out at the regional level. Colored rectangles refer to the timeframe when the different tiers were in place (see Table 1 in main text for a description of restrictions).

**Figure S4.**
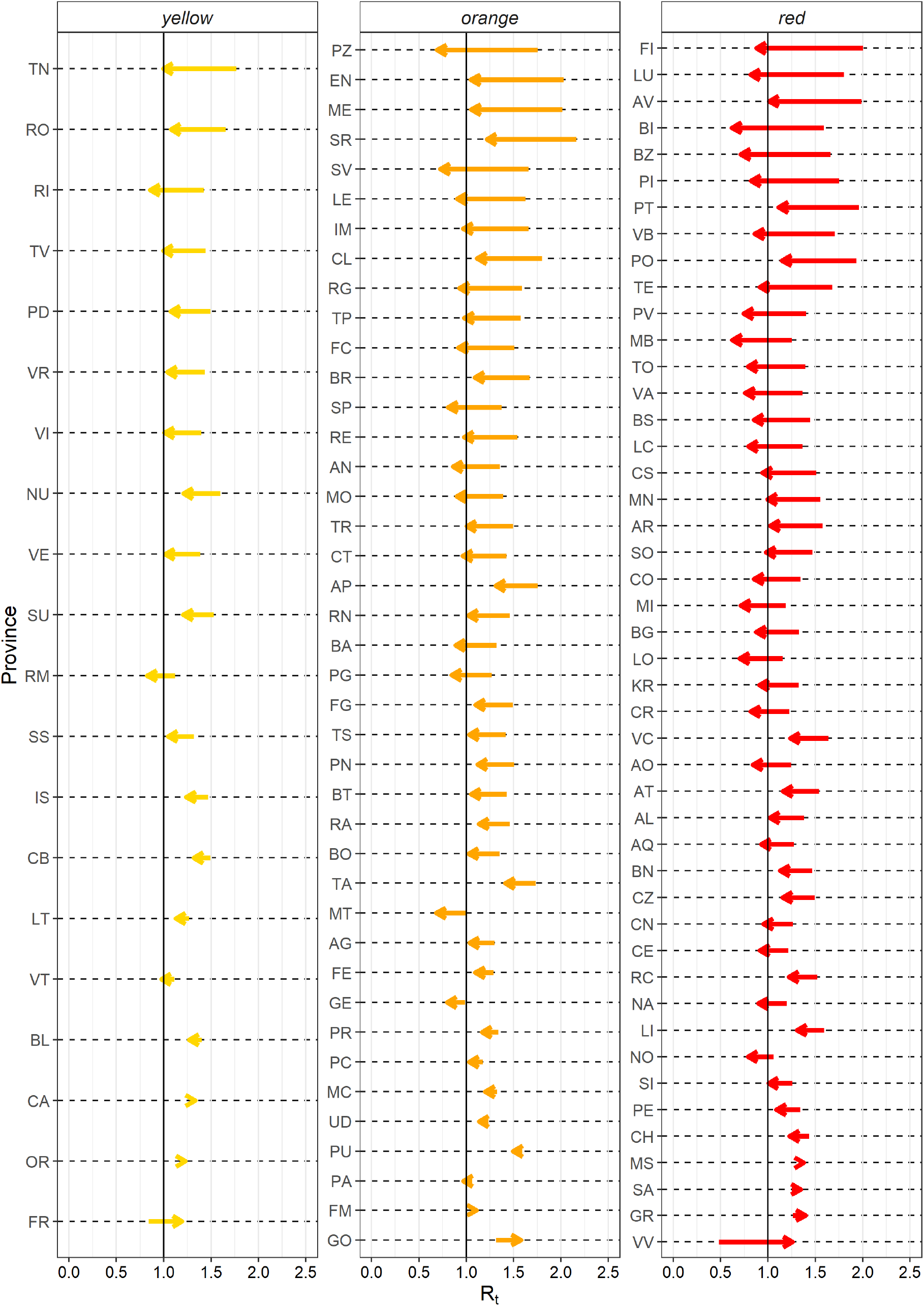
Variation in the net reproduction number from hospital admissions (Rt^h^) in each province. The arrows indicate the variation in Rt^h^ from the period before the introduction of the regional tiered restrictions (October 30-November 5) to the end of our observations (November 19 – 25). Provinces are ordered by decreasing reduction in Rt^h^.

**Table S6.** Result of the linear mixed model on the net reproduction number estimated from symptom onset, Rt, at the regional level. See Table S2 for interpretation of parameters.

| PARAMETERS | VALUE | STD<br>ERROR | DF | T-VALUE | P-VALUE |
| --- | --- | --- | --- | --- | --- |
| $\beta_0$ | 1.099 | 0.069 | 35.9 | 15.810 | <0.00001 |
| $\beta_1$ | 0.217 | 0.089 | 35.9 | 2.453 | 0.01914 |
| $\beta_2$ | 0.211 | 0.089 | 35.9 | 2.382 | 0.02265 |
| $\beta_3$ | -0.087 | 0.096 | 18 | -0.909 | 0.37562 |
| $\beta_4$ | -0.415 | 0.123 | 18 | -3.389 | 0.00327 |
| $\beta_5$ | -0.478 | 0.123 | 18 | -3.901 | 0.00105 |

**Table S7.** Mean net reproduction number estimated from symptom onset, Rt, at regional level before and after regional interventions.

| Maximum tier | Mean Rt (SD)<br>October 30 – November 5 | Mean Rt (SD)<br>November 19 - 25 | Relative<br>reduction |
| --- | --- | --- | --- |
| Yellow | 1.10 (0.13) | 1.01 (0.26) | 8.0% |
| Orange | 1.32 (0.19) | 0.81 (0.13) | 38.2% |
| Red | 1.31 (0.17) | 0.74 (0.09) | 43.2% |

**Figure S5.**
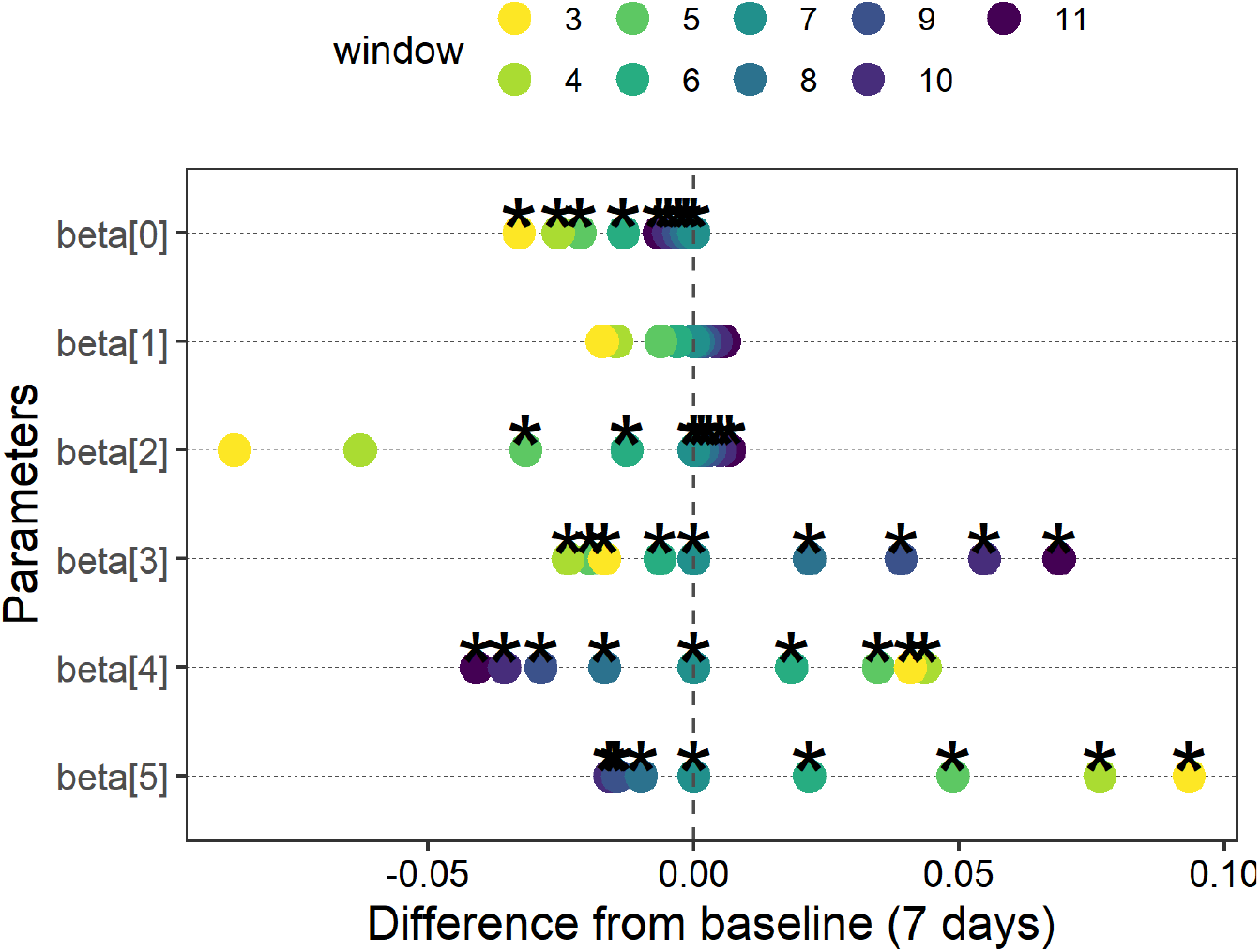
Variation in estimated model parameters when considering different lengths (in days) of the window over which Rt is averaged. On the x axis the difference between the estimated parameter value of the parameter and the main analysis (which uses a 7-day window). Parameter names are on the y axis. See Table S2 for interpretation of parameters. Colors represent different period lengths. Asterisks represent statistical significance of the estimated model parameters (p-value <0.05).

**Table S8.**
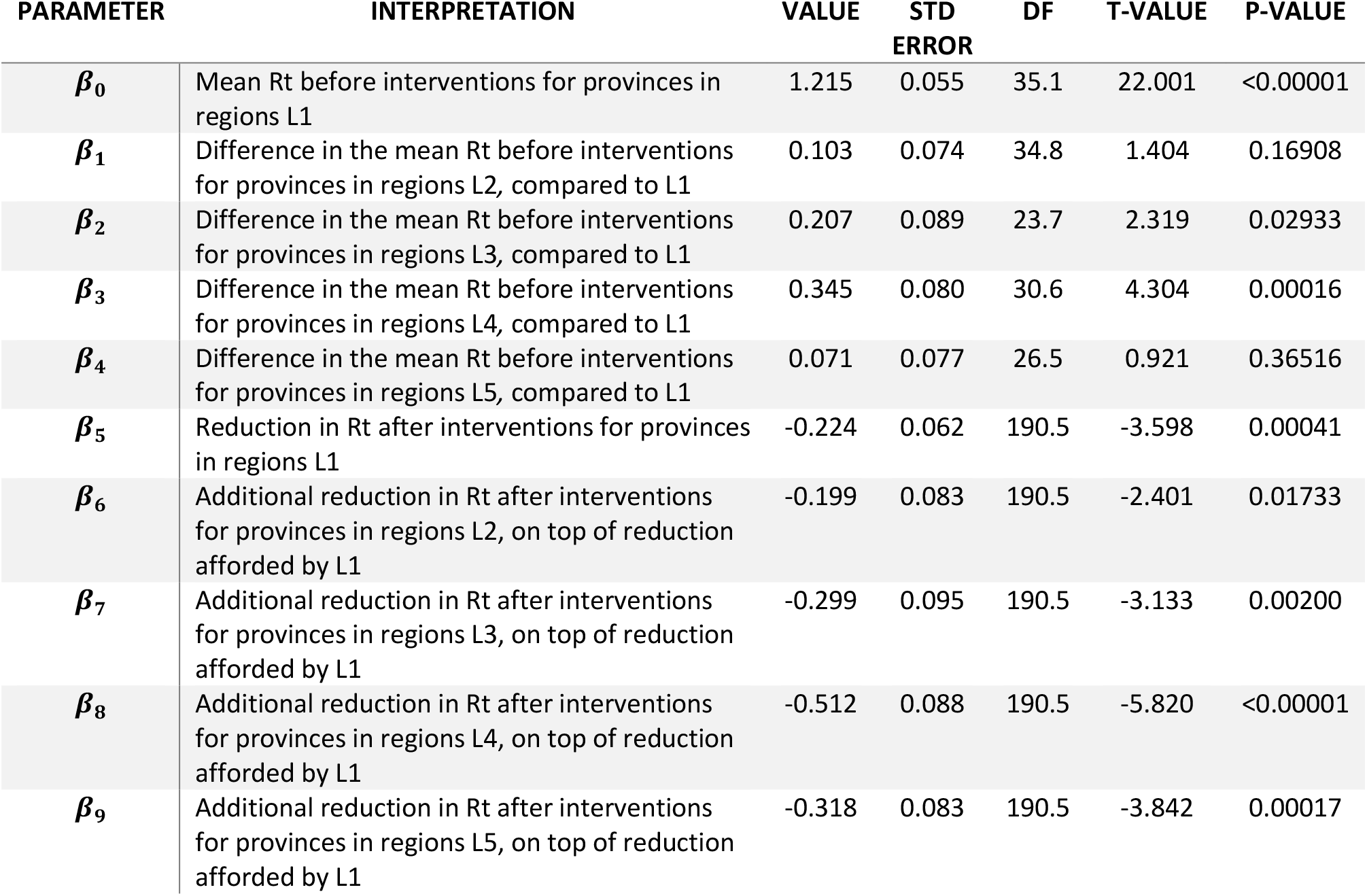
Result of the linear mixed model on the net reproduction number estimated from symptom onset Rt at provincial level considering 5 groups of interventions

**Table S9.** Mean net reproduction number estimated from symptom onset, Rt, before and after regional interventions.

| Tier | Mean Rt (SD)<br>October 30 – November 5 | Mean Rt (SD)<br>November 19 - 25 | Relative<br>reduction |
| --- | --- | --- | --- |
| L1 | 1.22 (0.23) | 0.99 (0.23) | 18.8% |
| L2 | 1.31 (0.25) | 0.89 (0.17) | 32.1% |
| L3 | 1.42 (0.22) | 0.90 (0.08) | 36.6% |
| L4 | 1.56 (0.26) | 0.82 (0.16) | 47.4% |
| L5 | 1.27 (0.24) | 0.73 (0.10) | 42.5% |

### 3. Impact on tiered restriction on hospital admissions

To estimate the impact of tiered-restrictions on the COVID-19 burden, we estimated the number of daily hospital admission expected under a scenario of constant transmissibility and compared these results with what expected by assuming a reduction in the transmission to Rt values in the different tiers. In the main analysis, simulations were obtained by applying the renewal equation to hospitalized cases, initialized at values observed over the window October 30 – November 5, 2020 using average estimates of Rt (Figure 4). As a sensitivity analysis with respect to this assumption, we projected the number of hospitalized cases by considering the variability in the estimate of the net reproduction number in each region. Specifically, we employed 1,000 different runs of the renewal equation (see Methods in the main text) where for each simulation we sampled an Rt value from a normal distribution of mean and variance equal to those characterizing the posterior distribution of Rt estimated from the time series of symptom onset for the period October 30 –November 5, 2020 (Figure S6). In this case, when assuming that the national restrictions existing on November 5 had been maintained until November 25, we estimated a cumulative incidence of hospital admissions of 75.7 (95%CI: 71.0 – 80.6), 107.6 (95%CI: 102.1 – 113.7) and 142.1 (95%CI: 137.0 – 147.3) per 100,000 in the yellow, orange and red tier respectively, resulting in a total of 70,193 (95%CI: 68,149-72, 990) hospital admissions (Figure S7). Maintaining national restrictions would have been resulted in a general increase of the daily hospital admission incidence between November 6 and November 25; for example, in the red tier regions/APs the incidence would have been risen from 5.3 per 100,000 per day on November 5 to 9.6 (95%CI: 9.0-10.2) per 100,000 per day on November 25 (Figure S8, Table S10). These values are in line with those observed in the main analysis and with limited variability around the mean estimates.

**Figure S6.**
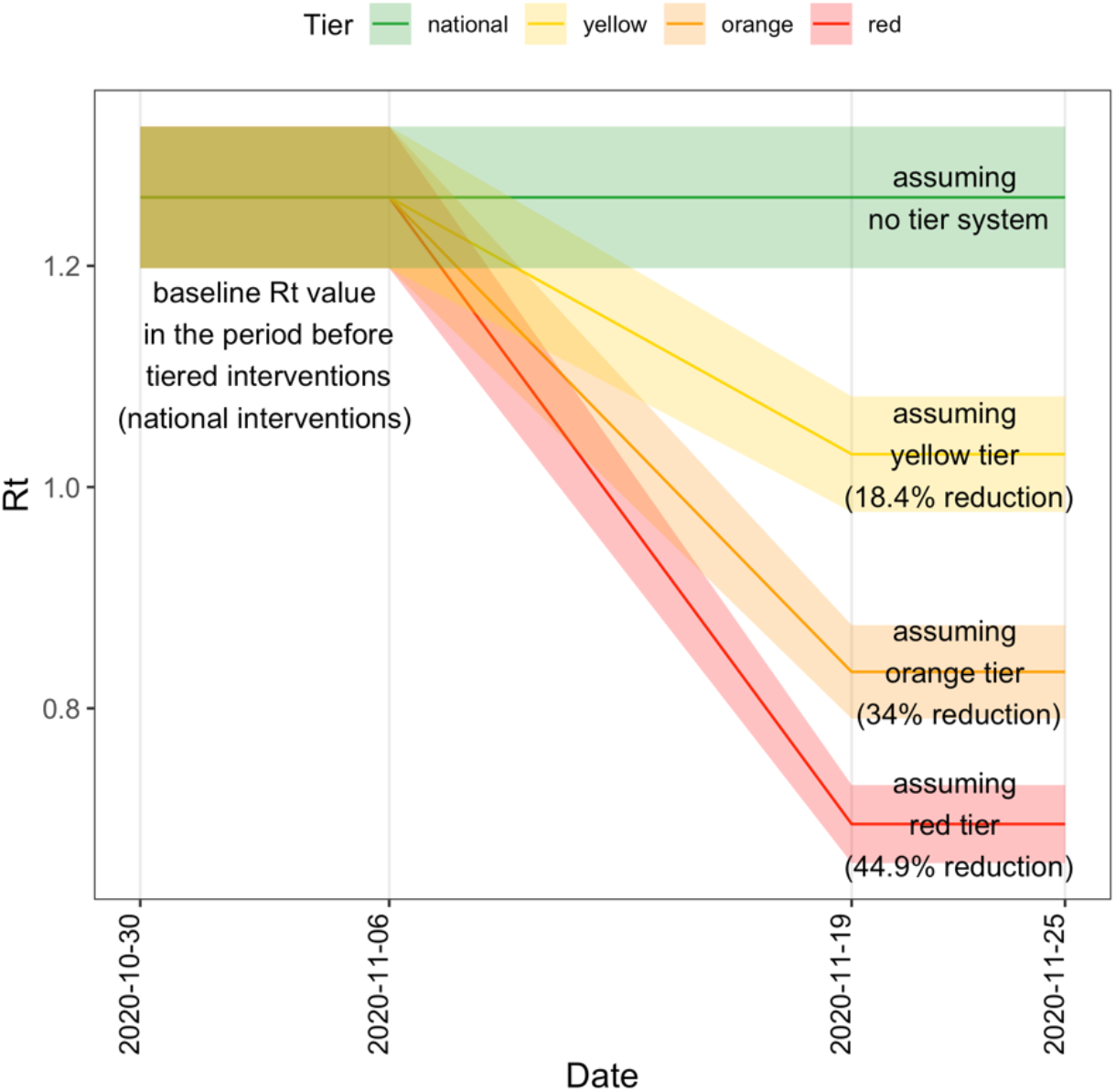
Schematic representation of the assumed profile of the net reproduction number under different intervention scenarios. Solid lines represent the mean Rt (included in the main analysis) while the shaded areas represent the variability around the mean Rt value (included in the sensitivity analysis).

**Figure S7.**
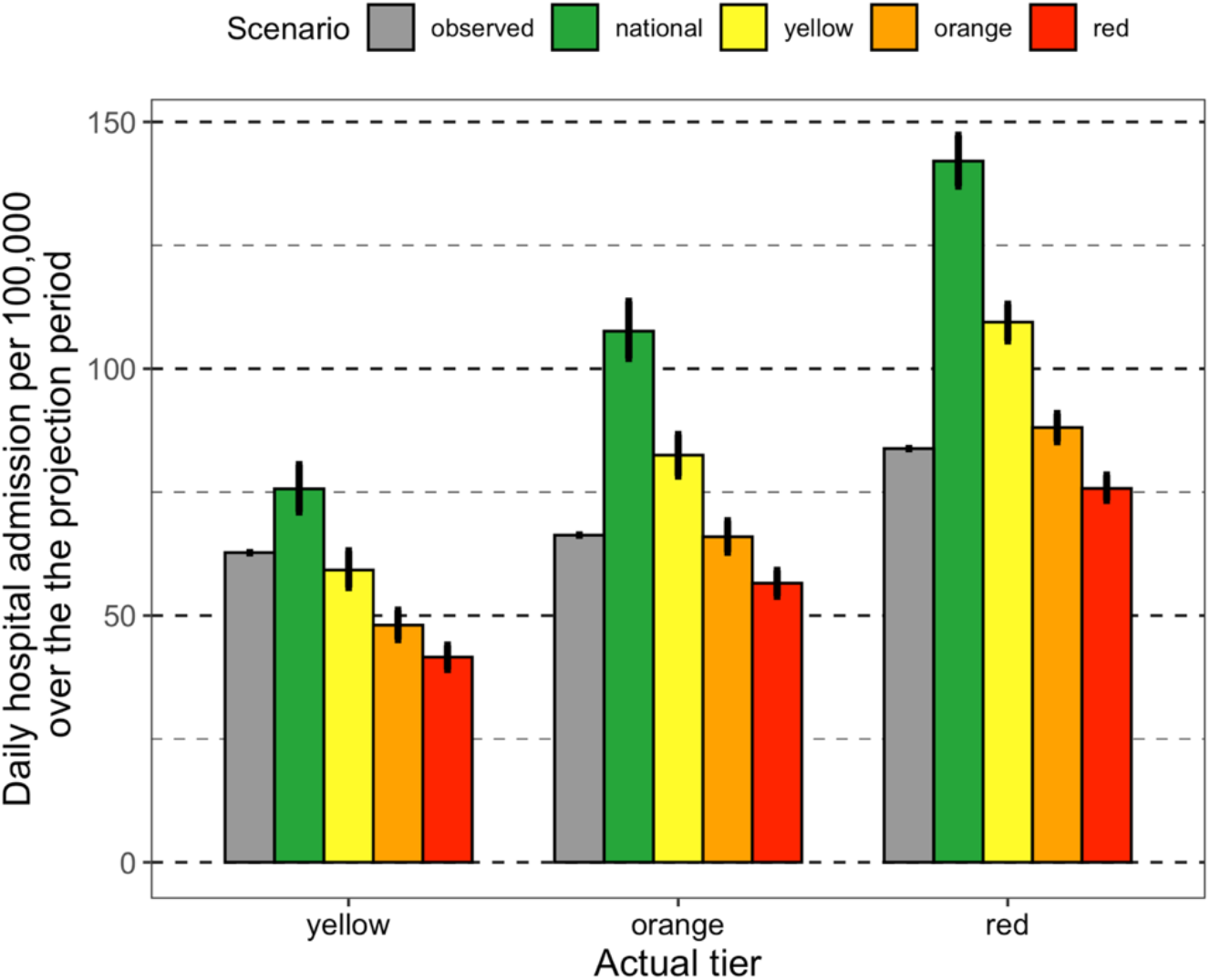
Incidence over the projection period (November 5 - November 25) in different scenarios. Gray bars represent the observed value for regions in different tiers. Colored bars represent the mean projected value under the assumption that restrictions are maintained over the study period. Vertical lines represent the 95% confidence interval of the projected distribution.

**Figure S8.**
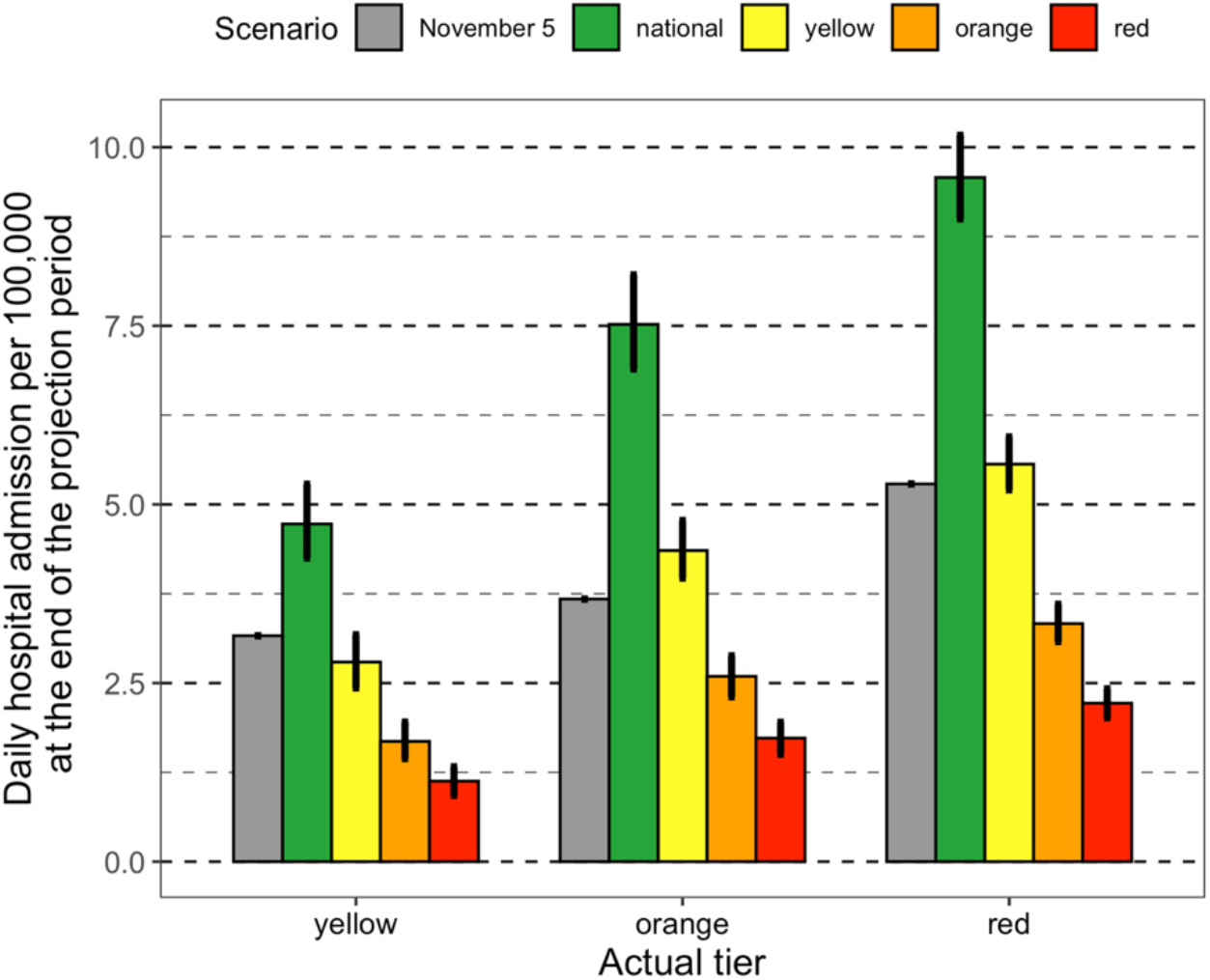
Incidence at the end of the projection period (November 25) in different scenarios. Gray bars represent the observed value at the beginning of the projection period (November 5) for regions in different tiers. Colored bars represent the mean projected value under the assumption that restrictions are maintained over the study period. Vertical lines represent the 95% confidence interval of the projected distribution.

**Table S10.** Scenario simulations for cumulative hospital admissions over the period November 6-25, 2020

| Actual tier | Observed hospital admissions | Population | Modeled tier | Mean projected hospital admissions (95%CI) | Mean projected hospital admissions (95%CI) [sensitivity] |
| --- | --- | --- | --- | --- | --- |
| **yellow** | 8,313 | 13,248,726 | national | 10,016 (9,450-10,552) | 10,025 (9,403-10,675) |
|  |  |  | yellow | 7,831 (7,422-8,235) | 7,846 (7,375-8,362) |
|  |  |  | orange | 6,362 (6,021-6,698) | 6,366 (5,978-6,771) |
|  |  |  | red | 5,507 (5,229-5,798) | 5,508 (5,176-5,830) |
| **orange** | 12,696 | 19,153,927 | national | 20,579 (19,758-21,425) | 20,609 (19,553-21,770) |
|  |  |  | yellow | 15,776 (15,124-164,35) | 15,798 (14,992-16,602) |
|  |  |  | orange | 12,626 (12,112-13,113) | 12,633 (12,036-13,255) |
|  |  |  | red | 10,816 (10,418-11,201) | 10,830 (10,326-11,335) |
| **red** | 23,341 | 27,843,345 | national | 39,468 (38,365-40,627) | 39,559 (38,142-41,010) |
|  |  |  | yellow | 30,461 (29,640-31,277) | 30,470 (29,410-31,487) |
|  |  |  | orange | 24,519 (23,786-25,244) | 24,524 (23,730-25,316) |
|  |  |  | red | 21,086 (20,546-21,692) | 21,095 (20,415-21,851) |

